# Adapting Lot Quality Assurance Sampling to accommodate imperfect tests: application to COVID-19 serosurveillance in Haiti

**DOI:** 10.1101/2020.09.11.20193052

**Authors:** Isabel R. Fulcher, Mary Clisbee, Wesler Lambert, Fernet Renand Leandre, Bethany Hedt-Gauthier

## Abstract

**Background:** Lot Quality Assurance Sampling (LQAS), a tool used for monitoring health indicators in low resource settings resulting in “high” or “low” classifications, assumes that determination of the trait of interest is perfect. This is often not true for diagnostic tests, with imperfect sensitivity and specificity. Here, we develop Lot Quality Assurance Sampling for Imperfect Tests (LQAS-IMP) to address this issue and apply it to a COVID-19 serosurveillance study in Haiti.

**Development:** As part of the standard LQAS procedure, the user specifies allowable classification errors for the system, which is defined by a sample size and decision rule. We show that when an imperfect diagnostic test is used, the classification errors are larger than specified. We derive a modified procedure, LQAS-IMP, that accounts for the sensitivity and specificity of a diagnostic test to yield correct classification errors.

**Application:** At Zanmi Lasante health facilities in Haiti, the goal was to assess the prior circulation of COVID-19 among healthcare workers (HCWs) using a limited number of antibody tests. As the COVID-19 antibody tests were known to have imperfect diagnostic accuracy, we used the LQAS-IMP procedure to define valid systems for sampling at eleven hospitals in Haiti.

**Conclusions:** The LQAS-IMP procedure accounts for imperfect sensitivity and specificity in system design; if the accuracy of a test is known, the use of LQAS-IMP extends LQAS to applications for indicators that are based on laboratory tests, such as COVID-19 antibodies.

**Key Messages:**

- It is imperative to account for the sensitivity and specificity of the diagnostic test in serosurveillance studies.
- For prevalence estimation and inference, adjustments for imperfect testing properties are available to researchers. However, no adjustments currently exist for the design of Lot Quality Assurance Sampling systems, resulting in invalid classification systems when used with an imperfect diagnostic test.
- When sensitivity and specificity of a diagnostic test is known, Lot Quality Assurance Sampling for Imperfect Tests should be used to determine the sample size and decision rule for a system as it will result in valid classification errors.

## Background

As the world scrambles to respond to the COVID-19 pandemic, timely and accurate surveillance is imperative to monitor the spread of the virus. SARS-CoV-2 virologic and serologic testing capacity is essential for individual clinical care, contact tracing and containment strategies, and monitoring for population-level dynamics. However, testing capacity in low- and middle-income countries (LMICs) is hindered by limited laboratory infrastructure and technicians, inability to compete on the global market to procure tests, absence of quality controls capability for the procured tests reagents, and limited resources to purchase the necessary tests [1]. Therefore, any testing activity in LMICs must be highly efficient, maximizing the information gained while minimizing the number of tests used.

Lot Quality Assurance Sampling (LQAS) is a classification procedure used to categorize an area as “high” or “low” on some trait of interest [2,3]. LQAS is used widely in monitoring and evaluation of health indicators in low resource settings [4,5] because the method produces results that can be quickly linked to program response, can be implemented without need for advanced statistical training or software, and because it often requires smaller sample sizes than traditional estimation procedures. An LQAS system is defined by a sample size and decision rule for classification, determined by user-specified definitions of high and low thresholds for performance and limits to the amount of classification errors tolerated at these thresholds.

A key assumption underlying the design of LQAS systems is that, for any individual sample, determination of the trait of interest is perfect. This is an unrealistic assumption in many settings, particularly for diagnostic tests where the occurrence of false positives and negatives is ubiquitous and often a known quantity. If the accuracy of the diagnostic test is not taken into account, the LQAS system can be deeply flawed resulting in sample sizes and decision rules that have higher classification errors than that specified by the user in the design. While methods that account for diagnostic testing accuracy have been well established for prevalence estimation [6,7], there is currently no method to account for imperfect diagnostic tests in the design of LQAS systems.

In this paper, we propose a new method for determining an LQAS system that accounts for the sensitivity and specificity of a diagnostic test. We present simulation studies to demonstrate the utility of our method and the impact of inaccurate estimates of sensitivity and specificity in various settings. We discuss the design of an LQAS system specific to the use of SARS-CoV-2 antibody tests among health care workers in Haiti as a motivating example for this method.

## Development

### Overview of Traditional LQAS Systems

In its most traditional form, the sample size, *n*, and decision rule, *d*, are determined by the population size, *N*, and then four parameters defined by users based on the specific context and goals. The four parameters are:

- *p_u_*, the upper prevalence threshold by which an area is classified as high;
- *p_ι_*, the lower prevalence threshold by which an area is classified as low; ¥
- *α*, the probability that a high area is mistakenly classified as low; and ¥
- *β*, the probability that a low area is mistakenly classified as high;

The sample size and decision rule are determined by finding the minimum *n* and corresponding *d*, such that the following are true:

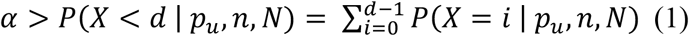

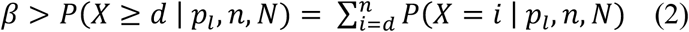

where *X* is a random variable denoting the number of persons with the trait of interest. Equation (1) states that the probability of sampling *n* individuals and observing fewer than *d* individuals with the trait of interest if the prevalence is *p_u_* and thereby erroneously classifying as “low” is less than *α* Equation (2) states that the probability of sampling *n* individuals and observing *d* or more individuals with the trait of interest if the prevalence is *p_ι_* and thereby erroneously classifying as “high” is less than *β*.

If working with small population sizes, for example facilities with a small number of health care workers, it is advantageous to take into account the population size, *N*, in these calculations as smaller population sizes will require a lower sample size. For small population sizes, the above probabilities are calculated using the hypergeometric distribution. That is, *X* ∼ *HGM*(*N,n,p_i_*), where *i* corresponds to the lower or upper prevalence threshold. If you have a sufficiently large population, then *N* is not necessary and the above probabilities can be calculated using the binomial distribution for *X*. There are freely available online tools and software packages to calculate the LQAS system based on both scenarios [8,9].

### Accounting for imperfect testing properties

A problem with the traditional approach is that it assumes the method used to identify a “case” is perfect. That is, all positives are true positives and all negatives are true negatives. However, the majority of diagnostic tests suffer from imperfect diagnostic capabilities characterized by test sensitivity (*S_e_*) and specificity (*S_p_*). This is certainly the case with COVID-19 antibody tests developed during the first months of the pandemic. Therefore, there is a possibility that *X* includes some false positives and that some true positives are not counted in *X*.

We propose the following adjustment to Equations (1) and (2),

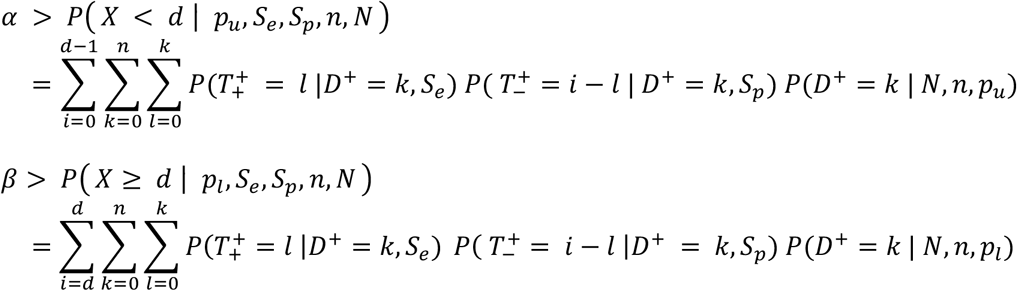

where *D*^+^, denotes the number of diseased persons chosen in the sample of size *n* and has a hypergeometric distribution with parameters *N, n*, and *p_i_* (*i* corresponds to the lower or upper prevalence thresholds). Let 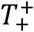 denote the number of true positives given by a binomial distribution with parameters *D*^+^ and *S_e_* and 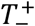 denote the number of false positives given by a binomial distribution with parameters *n* – *D*^+^ and 1 – *S_p_*. The proof is given in the Appendix.

We refer to this modified system as LQAS-IMP (<u>L</u>ot <u>Q</u>uality <u>A</u>ssurance <u>S</u>ampling for <u>Imp</u>erfect tests). These adjustments are important to reflect the true errors of misclassification. Once they are made, *α* and *β* will still account for uncertainty due to sampling (as is typical in LQAS), but will now also account for the uncertainty due to using imperfect tests. These modified equations reduce to Equations (1) and (2) under a perfect diagnostic test with sensitivity and specificity equal to one. The code for LQAS-IMP is available at: https://github.com/isabelfulcher/lqas_imp.

We make one cautionary note. It may be tempting to simply adjust the upper and lower prevalence thresholds to account for sensitivity and specificity of the test, that is substituting *p_i_*\* as defined below for *p_i_* in Equations (1) and (2),

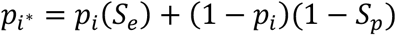

This will result in invalid *α* and *β* classification errors as this implicitly assumes that the sensitivity and specificity correspond to a fixed number of false positives and false negatives in a sample. Unlike the upper and lower prevalence thresholds, these probabilities cannot be considered fixed values for a given sample – for each individual tested, the probability of a false positive or false negative is drawn from a binomial distribution that depends on *S_e_* and *S_p_* – whereas the underlying disease status is fixed for that individual at the outset.

## Application

### Application to serosurveillance in Haiti

At Partners In Health in Haiti, known locally as Zanmi Lasante, the goal was to assess the prior circulation of COVID-19 among healthcare workers (HCWs) at eleven facilities of varying sizes. HCWs have higher exposure risks than the general population, both in their clinical care and support activities and because they typically move more between home and facility locations. Therefore, HCWs can be monitored to detect any early evidence of disease circulation. If there is little-to-no evidence of COVID-19 antibodies in this high-risk population, then it is unlikely that SARS-CoV-2 is circulating or has circulated in the general population. Evidence of high antibody prevalence gives indication for the high-risk population’s exposure that can be followed with additional studies of population transmission dynamics.

Like other programs in LMICs, Zanmi Lasante has limited testing resources and needs to balance the resources used for this surveillance activity with other monitoring activities that require rapid antibody tests. For that reason, the team at Zanmi Lasante proposed designing a Lot Quality Assurance Sampling (LQAS) approach to classify facilities as having high or low COVID-19 antibody prevalence in HCWs. The available tests were antibody (IgG/IgM) rapid diagnostic tests. The ZL team originally estimated the sensitivity and specificity to be 90%, based on PIH synthesis of manufacturers details and other reports on test properties.

The Haiti team converged on a final set of parameters that all team members agreed met the overall program goals and aligned with their plans for next steps after the high and low classifications. Based on the Haiti team’s extensive knowledge of their health facilities and patient population, the parameter values chosen were *p_u_* = 0.15, *p_ι_* = 0.05, and *α* = *β* = 0.10. That is, a “high prevalence” hospital is one where greater than or equal to 15% of workers test positive for antibodies and a “low prevalence” hospital is one where less than or equal to 5% of workers test negative for antibodies. The probability that a truly high prevalence hospital is mistakenly classified as low is 10%, and the probability that a truly low prevalence is mistakenly classified as high is 10%.

We note that the challenge with defining these parameters is that there is no inherently “right” or “wrong” value choice. Different programs will choose different values depending on their *a priori* knowledge of underlying prevalence levels and to overall goals and activities of that program. For instance, some programs may consider certain errors worse than others. Parameters may be set to either find more problem areas or, more often in the case of limited resources, set to only find the worst of the worst.

The sample sizes and decision rules for each health facility under our approach are given in Table 1. Importantly, for Belladere, there were no values of *n* and *d* that satisfied the *α* and *β* constraints, so we chose values that minimized both of these errors (see Simulation Study section). Under a perfect test (sensitivity and specificity both equal to one), the required sample size and decision rule would be *n* = 60 and *d* = 6 for Mirebalais (*N* = 1373). As such, the required sample size more than doubles to account for diagnostic accuracy of the test. For prevalence estimation with a corresponding 95% confidence interval of length 10% (+/−5%), the sample size required would be *n* = 172 (assuming an underlying prevalence of 15%, perfect test accuracy, and N = 1373). This increases to n = 251, a 68% increase in sample size, for prevalence estimate with a corresponding confidence interval of +/−5%, when accounting for an imperfect test sensitivity and specificity of 90%.

**Table 1:** LQAS-IMP system accounting for 90% sensitivity and specificity of the antibody tests for eleven health facilities in Haiti (*α* = *β* = 0.10, *p_u_* = 0.15, *p_l_* = 0.05)

| **Hospital** | **Population, $N$** | **Sample size, $n$** | **Decision rule, $d$** |
| --- | --- | --- | --- |
| Mirebalais | 1373 | 149 | 27 |
| Cange Campus | 655 | 144 | 26 |
| Sant Marc | 533 | 143 | 26 |
| Hinche | 228 | 121 | 22 |
| Petite Riviere | 199 | 120 | 22 |
| Lascahobas | 184 | 109 | 20 |
| Verettes | 130 | 98 | 18 |
| Thomond | 124 | 97 | 18 |
| Cerca La Source | 123 | 109 | 20 |
| Belladere | 110 | 108 | 20 |
| Boucan Carre | 108 | 98 | 18 |

### Method performance and robustness to misspecified test properties: A simulation study

#### Comparison of LQAS and LQAS-IMP for Haiti example

We validated our proposed approach and demonstrated the impact of adjusting for incorrect sensitivity and specificity of a test in a simulation study. To validate our approach, we calculated the *α* and *β* classification errors under standard LQAS and LQAS-IMP for all hospitals in the Haiti example. Under standard LQAS, which implicitly assumes *S*_*e*_ = *S*_*p*_ = 1, we calculated the sample size and decision rule for each hospital (Table 2). For each hospital, we then generated 3000 datasets of size *N* with a) the true prevalence set at the probability threshold (*p*_*u*_ for *α*; *p*_*ι*_ for *β*) to generate the true antibody status for a sampled individual and b) test sensitivity and specificity of 90% to generate the observed test result for each person given their antibody status. We then computed *α* and *β* using the sample sizes and decision rules from both Table 1 (adjusted) and Table 2 (standard) for comparison.

**Table 2:**
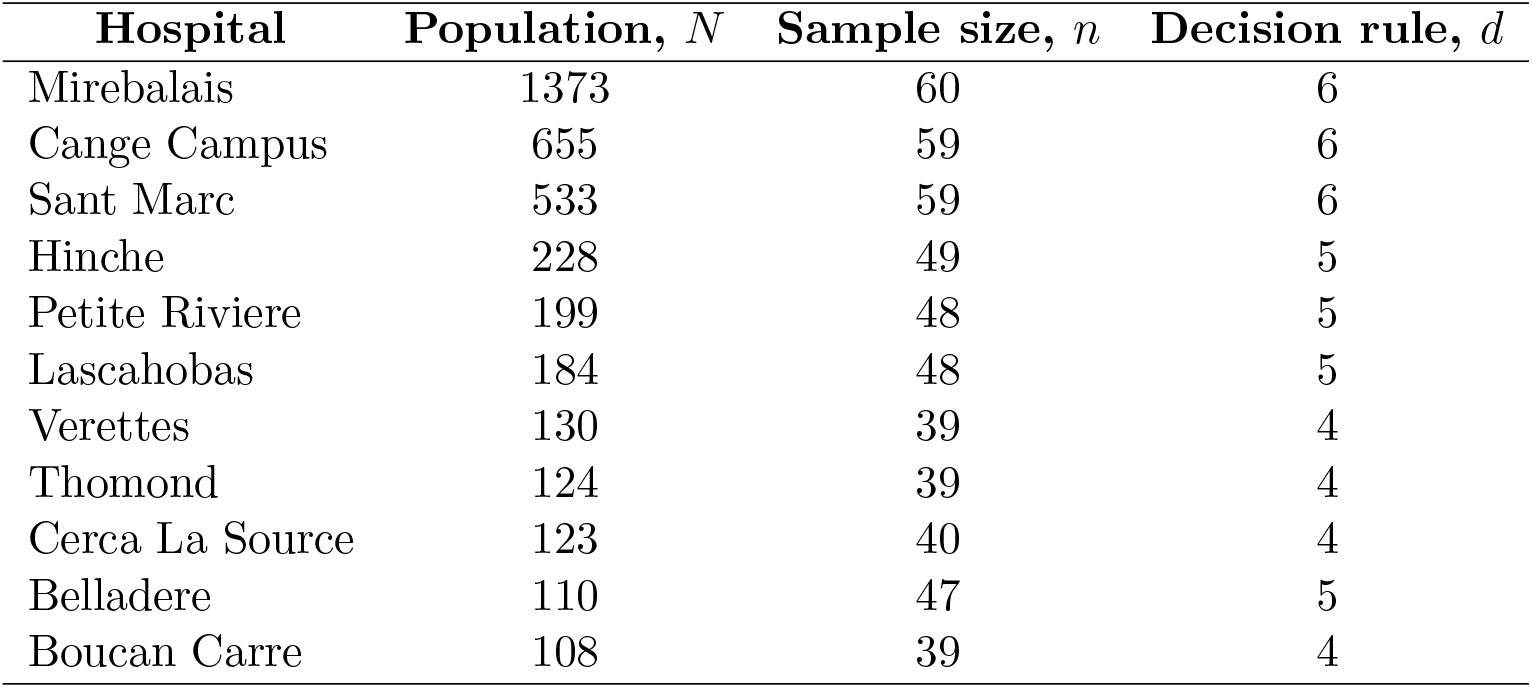
Standard LQAS system assuming perfect antibody test accuracy for eleven health facilities in Haiti (*α* = *β* = 0.10, *p_u_* = 0.15, *p_l_* = 0.05)

Unsurprisingly, the standard LQAS required substantially smaller sample sizes than LQAS-IMP. Figure 1 shows the observed *α* and *β* errors across the simulated datasets for each hospital.

**Figure 1:**
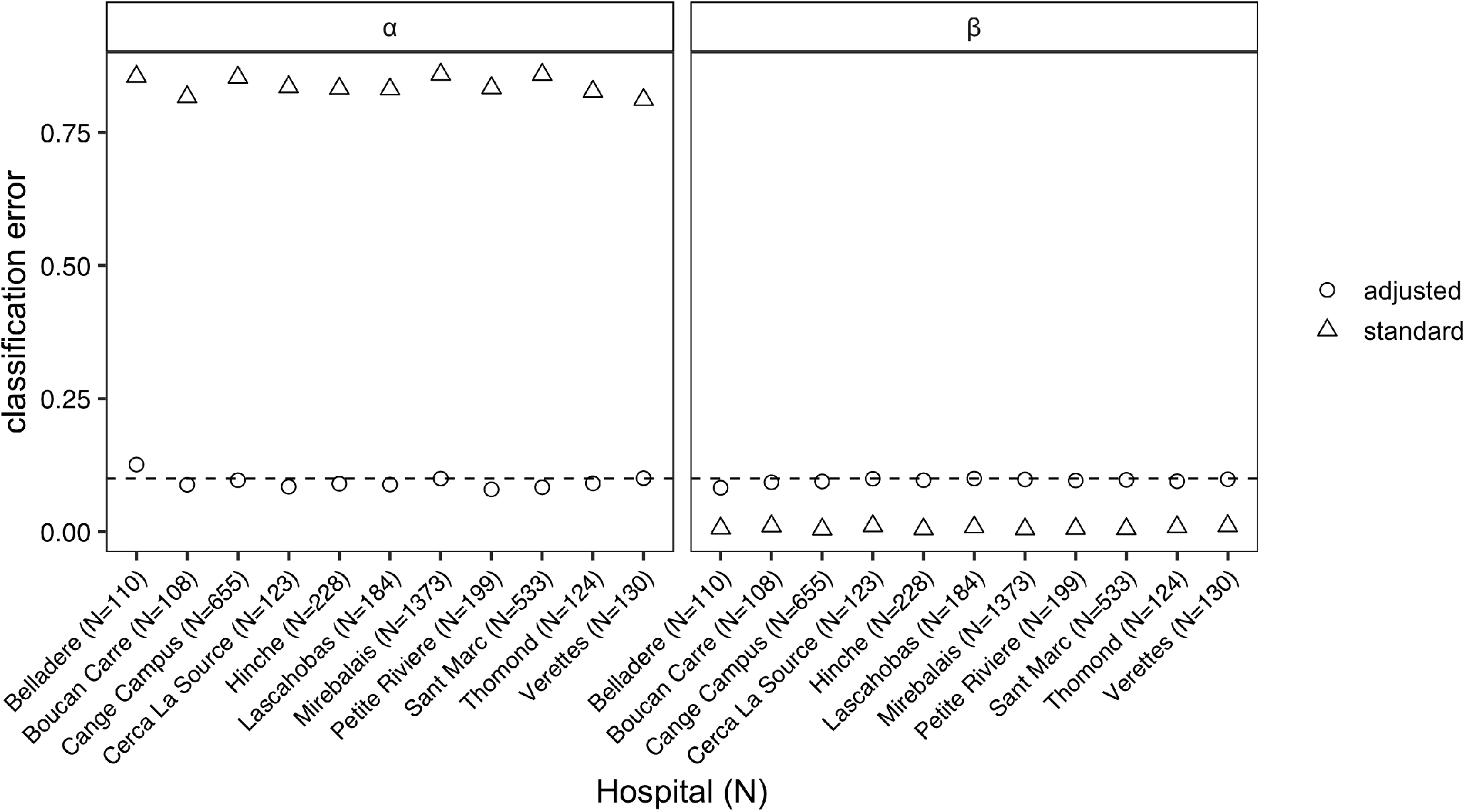
The classification errors for each hospital from LQAS (standard) and LQAS-IMP (adjusted) classification systems when test sensitivity and specificity is equal to 90% (*p_u_* = 0.15 and *p_l_* = 0.05).

When the sensitivity and specificity are 90%, using the sample size and decision rule from the standard approach (that does not account for test properties) results in substantially higher *α* values than desired (ranging from 0.81-0.86 vs. 0.10). For example, the standard LQAS system resulted in 86% of the high antibody prevalence (*p*_*u*_ = 0.15) datasets for Mirebalais being incorrectly classified as low prevalence. In contrast, the standard approach resulted in much lower *β* values that specified (0.01 for all hospitals vs. 0.10). The behavior of the classification errors are due to the low prevalence for *p_l_* and *p_u_*, and if both were closer to one than we would observe overly high *β* values and lower *α* values.

LQAS-IMP demonstrated *α* and *β* values consistent with the targeted maximum classification error of 0.10. The one exception was for Belladere, as our method could not find a system that satisfied the targeted classification error. The best system for this hospital resulted in *α* = 0.13 and *β* = 0.08 as shown in Figure 1.

#### Robustness of classification errors to misspecified sensitivity and specificity in LQAS-IMP

We also varied the true test sensitivity and specificity from 0.85 to 0.95 to investigate changes in *α* and *β* classification errors when the actual test sensitivity and specificity is different than what was specified in the LQAS-IMP design. We conducted the simulation for three upper and lower prevalence thresholds: low (*p*_*u*_ = 0.15 and *p*_*ι*_ = 0.05), medium (*p*_*u*_ = 0.55 and *p*_*ι*_ = 0.45), and high (*p*_*u*_ = 0.95 and *p*_*ι*_ = 0.85). For the three prevalence thresholds, we calculate the required *n* and *d* with LQAS-IMP accounting for sensitivity and specificity equal to 0.9 and *α* and *β* error equal to 0.10, as in the Haiti setting. We computed the estimated *α* and *β* from 3000 generated datasets of size *N* = 228 with a true prevalence set at the prevalence threshold (*p*_*u*_ for *α*; *p*_ι_ for *β*) and varied sensitivity and specificity. The LQAS-IMP system was *n* = 121 and *d* = 18 for the low probability threshold, *n* = 157 and *d* = 79 for the medium probability threshold, and *n* = 121 and *d* = 100 for the high probability threshold.

For the low prevalence thresholds, an incorrect assumed specificity greatly impacted the *α* and *β* misclassification errors (Figure 2A). That is, if one assumed the specificity was 90% in the LQAS-IMP design, but it was truly 89%, the α error would be 16% rather than the target 10%. However in this scenario, an incorrect assumed sensitivity has minor impact on the classification errors.

For the high probability threshold (Figure 2C), the opposite held – an incorrect assumed sensitivity impacts the *α* and *β α* misclassification errors, whereas an incorrect sensitivity has only minor impact. For the medium probability threshold, incorrect assumed specificity or sensitivity have major impacts on both classification errors.

**Figure 2:**
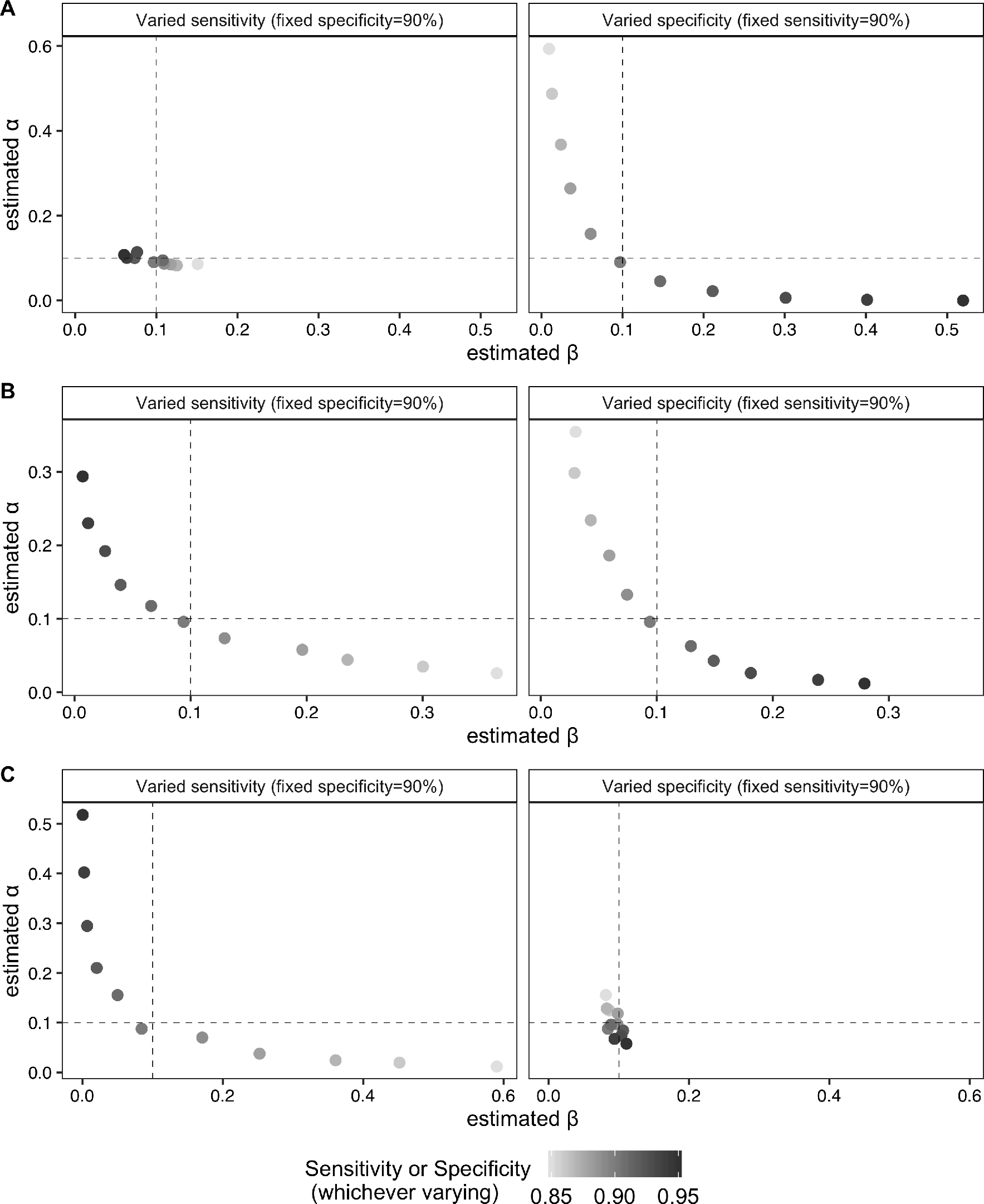
Misclassification errors for varying values of true sensitivity and specificity under (A) low, (B) medium, (C) high prevalence thresholds. The assumed sensitivity and specificity for adjustment was 90%. The dotted lines indicate the pre-determined values of *α* and *β*. The black dot represents alignment between the true and assumed sensitivity and specificity values, which should always be contained in lower left square.

## Discussion

### LQAS-IMP in the context of serosurveys

A requirement of the LQAS-IMP system is that test sensitivity and specificity are known and fixed quantities. Although estimates of sensitivity and specificity are often “known” as they are reported with diagnostic tests [10,11], there may be variability in these estimates due to finite sample sizes in clinical agreement studies. For example, in the early months of COVID-19, the sample sizes for antibody test agreement studies was quite small (< 200 samples), resulting in wide confidence intervals for test sensitivity and specificity [12]. As shown in the simulation study, small deviations from the assumed sensitivity or specificity in LQAS-IMP can result in larger than expected misclassification probabilities. We encourage researchers to conduct similar simulation studies to assess the potential impact of incorrect sensitivity and specificity estimates on their systems. In the case of unknown or unreliable estimates of sensitivity and specificity, LQAS-IMP may not be suitable and prevalence estimation with appropriate adjustment is the best path forward [6,7]. Ultimately, after evaluating the performance of LQAS-IMP and assessing our uncertainty of the COVID-19 antibody test properties, we decided that this approach was not appropriate for the Zanmi Lasante context. However, as these tests and our confidence in the test properties improve, this may change.

### Combining multiple LQAS-IMP procedures for prevalence estimation

As noted throughout, LQAS is a classification procedure. However, the results can also be used for estimation as the *n* individuals are a random sample [13]. Typically, estimation at the same level as classification is discouraged because the sample sizes are generally small resulting in confidence intervals that are too wide to be informative. In the Haiti case, there are multiple sites and combining samples across sites could produce overall estimates of antibody prevalence with high precision. The estimation process is straightforward; prevalence and variance estimates can be calculated from basic formulae from stratified sampling theory using standard software. However, the final prevalence and variance estimates must account for the imperfect sensitivity and specificity of the tests. These methods are well-defined [6], with recent extensions to COVID-19 prevalence estimates [7].

## Conclusion

The LQAS-IMP procedure allows teams to design assessments to extend the advantage of the positive features of LQAS to measures based on tests with imperfect properties. We originally designed this method to support Zanmi Lasante’s COVID-19 serosurveys. As we assessed the method’s properties, and specifically the vulnerability to misspecified sensitivity and specificity, we ultimately decided it was premature to deploy this method for this purpose. However, as COVID-19 antibody tests improve and the properties better quantified, LQAS-IMP may provide a rapid way to assess the state of COVID-19 burden in a context. This method can also be readily applied to monitoring other diseases.

## Data Availability

Not applicable

https://github.com/isabelfulcher/lqas_imp

## Acknowledgements

We thank the Zanmi Lasante team for their helpful input on this study, specifically, Cynthia Charles (Nurse Epidemiologist) and Betty Alexandre Pierre (Zanmi Lasante Lab Coordinator). Bethany Hedt-Gauthier received funding from the Global Health Research Core at Harvard Medical School for this research.

## References

[1] Kavanagh MM, Erondu NA, Tomori O, Dzau VJ, Okiro EA, Maleche A, Aniebo IC, Rugege U, Holmes CB, Gostin LO. Access to lifesaving medical resources for African countries: COVID-19 testing and response, ethics, and politics. The Lancet 2020;395:1735–1738.

[2] Dodge HF and Romig HG. A method of sampling inspection. The Bell System Technical Journal 1924;8:613–631.

[3] Shewhart WA. Economic control of quality of manufactured product. London: Macmillan And Co Ltd, 1931.

[4] Robertson SE and Valadez JJ. Global review of health care surveys using lot quality assurance sampling (LQAS), 1984-2004. Social science & medicine 2006;63(6):1648–1660.

[5] Lanata C, Stroh G, Black R. Lot quality assurance sampling in health monitoring. The Lancet 1988;331:122–123.

[6] Diggle PJ. Estimating prevalence using an imperfect test. Epidemiology Research International 2011.

[7] Larremore DB, Fosdick BK, Bubar KM, Zhang S, Kissler SM, Metcalf CJE, Buckee C, Grad Y. Estimating SARS-CoV-2 seroprevalence and epidemiological parameters with uncertainty from serological surveys. medRxiv 2020.

[8] Office of Health, Infectious Diseases, and Nutrition, Bureau for Global Health, United States Agency for International Development. http://www.brixtonhealth.com/hyperLQAS.html (August 2020, date last accessed).

[9] Valadez J. http://lqas.spectraanalytics.com/ (August 2020, date last accessed).

[10] U.S. Department of Health and Human Services, Food and Drug Administration, Center for Devices and Radiological Health. Policy for Coronavirus Disease-2019 Tests During the Public Health Emergency (Revised). 2020. https://www.fda.gov/media/135659/download (August 2020, date last accessed).

[11] United States Food and Drug Administration. EUA Authorized Serology Test Performance. 2020. https://www.fda.gov/medical-devices/coronavirus-disease-2019-covid-19-emergency-useauthorizations-medical-devices/eua-authorized-serology-test-performance (August 2020, date last accessed).

[12] Center for Health Security, Johns Hopkins University. Serology-based tests for COVID-19. 2020. https://www.centerforhealthsecurity.org/resources/COVID-19/serology/Serology-basedtests-for-COVID-19.html#sec1 (August 2020, date last accessed).

[13] Olives C, Valadez JJ, Pagano M. Estimation after classification using lot quality assurance sampling: corrections for curtailed sampling with application to evaluating polio vaccination campaigns. Tropical Medicine & International Health 2014;19(3):321–330.

